# Parental and individual socioeconomic position show distinct associations with trajectories of diet quality across adolescence and early adulthood

**DOI:** 10.1101/2024.11.13.24317238

**Authors:** Oscar Rousham, Tanya Braune, Trevor A Mori, Lawrence J Beilin, Eleanor M Winpenny

**Affiliations:** Sheffield Centre for Health and Related Research (SCHARR), The University of Sheffield, Sheffield, S10 2TN, UK; Mohn Centre for Children’s Health and Wellbeing, School of Public Health Imperial College London, London, W2 1PG, UK; MRC Epidemiology Unit, The University of Cambridge, Cambridge, CB2 0SL, UK; Medical school, The University of Western Australia, Perth, WA, Australia

## Abstract

**Background:** Adolescence and early adulthood is an important period for the development of health behaviours and diet quality, which may persist through adult life. Socioeconomic position (SEP) during early adulthood shows associations with inequalities in diet quality and cardiometabolic health later in adulthood, yet little is known about how dietary inequalities develop during adolescence and early adulthood. This study aimed to identify distinct diet quality trajectories observed across adolescence and early adulthood; and explore the association between parental and individual SEP and these trajectories.

**Methods:** The Raine Study is a large multigenerational cohort (n=1984) based in Western Australia. Dietary data was self-reported at five time points from ages 14 to 27 years. Growth mixture modelling was used to identify distinct diet quality trajectories based on Dietary Approaches to Stop Hypertension (DASH) diet scores. Multinomial logistic regression was used to determine the associations of parental SEP (maternal education level; and household income at age 14) and individual SEP (school leaving age) with diet quality trajectory membership.

**Results:** Three diet quality trajectories were identified: consistently low quality (41%), low-to-high quality (42%), and consistently high quality (16%). The ‘low-to-high’ diet quality trajectory showed a steep increase in diet quality from ages 14 to 20 years. Higher parental SEP was associated with a higher diet quality trajectory. After adjusting for parental SEP, leaving school before year 11 remained strongly associated with the low diet quality trajectory.

**Conclusion:** Our study demonstrates the development of dietary inequalities across adolescence and early adulthood, highlighting the importance of emerging individual SEP, in addition to parental SEP in shaping this. Interventions to improve diet quality in early adulthood are particularly needed among those who leave education at a young age.

## 1 Background

Diet quality is a key risk factor for chronic diseases (Ruiz et al., 2019). At the food group level, some food groups, such as fruit and vegetables, have been shown to be protective against chronic disease (He et al., 2007). In contrast, high consumption of other food groups, such as sugar-sweetened beverages, have been associated with an increased risk of chronic diseases like type 2 diabetes, obesity and CVD (Malik et al., 2010; Malik and Hu, 2022). Meanwhile, trials of healthy diets, such as the DASH diet or the Mediterranean Diet, have led to improvements in blood pressure (Appel Lawrence J. et al., 1997) and a decreased risk of cardiovascular disease (Estruch et al., 2018) and diabetes (Kargin, Tomaino and Serra-Majem, 2019). Diet is not fixed, but evolves over time, with food choices shaped by both current and lifetime socioenvironmental context and experiences with food. As a result, diets can be understood as trajectories which develop over time (Devine, 2005; Chong, 2022).

Adolescence and early adulthood are critical periods during which inequalities in diet quality may develop. During this period diet quality has been found to be poor and the prevalence of overweight and obesity increases sharply (Johnson et al., 2015; Winpenny et al., 2018). As of 2017, 46% of 18-24 year olds living in Australia were overweight or obese (Overweight and obesity, Summary, 2023). Adolescence and early adulthood are characterised by social transitions and changing socioenvironmental influences such as leaving school, entering the workplace and developing new peer networks. These environmental factors are known to influence food choices (Devine, 2005; Munt, Partridge and Allman-Farinelli, 2017; Winpenny et al., 2018) and diet quality and the consumption of specific food groups (e.g. fruits and vegetables, and sugar sweetened beverages) have been shown to change markedly during this period (Hiza et al., 2013; Banfield et al., 2016; Winpenny et al., 2017; Winpenny et al., 2018, p. 1). Adolescence and early adulthood are an opportune period for dietary interventions which could exploit the habit-breaking nature of social transitions to achieve long-term improvements in diet quality (Verplanken and Roy, 2016).

Inequalities in diet quality in adulthood according to socioeconomic and sociodemographic characteristics have been well established (Darmon and Drewnowski, 2008, 2015). Cross-sectional studies have shown the importance of parental SEP, in particular maternal education level (Desbouys et al., 2020), in shaping diet quality during adolescence and early adulthood. Other studies have shown the association between individual occupation and education level, and diet quality during this period (Olinto et al., 2011; Desbouys et al., 2019; Sexton-Dhamu et al., 2021). Further, a study demonstrated the effect of individual education level, independent of parental education level, on diet quality during this period (Faught et al., 2019). These studies are limited to a single wave of dietary data and do not capture an individual’s changing diet quality during adolescence and early adulthood. A small number of studies have applied latent trajectory analysis to longitudinal dietary data during adolescence and early adulthood. These have shown associations between diet quality trajectories and parental socioeconomic status (Dalrymple et al., 2022) and individual sex (Doggui et al., 2021). One study found a positive association between diet quality trajectories during this period and maternal education level using the Raine Study dataset (Appannah et al., 2021). However, these studies do not consider how the influence of individual SEP, in addition to parental SEP, begins to contribute to inequalities in diet quality over this period.

In this study we aimed to answer three research questions: 1. What are the different diet quality trajectories identified across adolescence and early adulthood? 2. What food groups drive the difference in diet quality between these trajectories? 3. How are parental and individual socioeconomic position associated with following different diet quality trajectories?

## 2 Methods

### 2.1 Cohort design and study population

This study used data from Generation 2 (Gen2) of the Raine Study, a longitudinal cohort study based in Perth, Western Australia. The study cohort have been described in detail elsewhere (White et al., 2017). In brief, Gen2 of the Raine Study began with 2868 participants born in 1989-1991. Dietary data have been collected at mean ages 14,17,20, 22 and 27 years for a total of 1984 participants.

Participants were included in this analysis if diet data was available for at least one of these five time points. Participants were excluded from a wave if: they had incomplete FFQ data; they were pregnant, to account for the change in nutritional requirements during pregnancy (Eckl, Brouwer-Brolsma and Küpers, 2021); or their total daily energy intake was unfeasibly high (>1.5 times estimated required calories) or low (<500Kcal for females, <800Kcal for males) (Willett, 2012). Accounting for increased energy requirements during adolescence (Scientific Advisory Committee on Nutrition, 2012) this gives a plausible daily energy intake range of 800-4800Kcal in males and 500-3600Kcal in females (Figure 1).

**Fig. 1.**
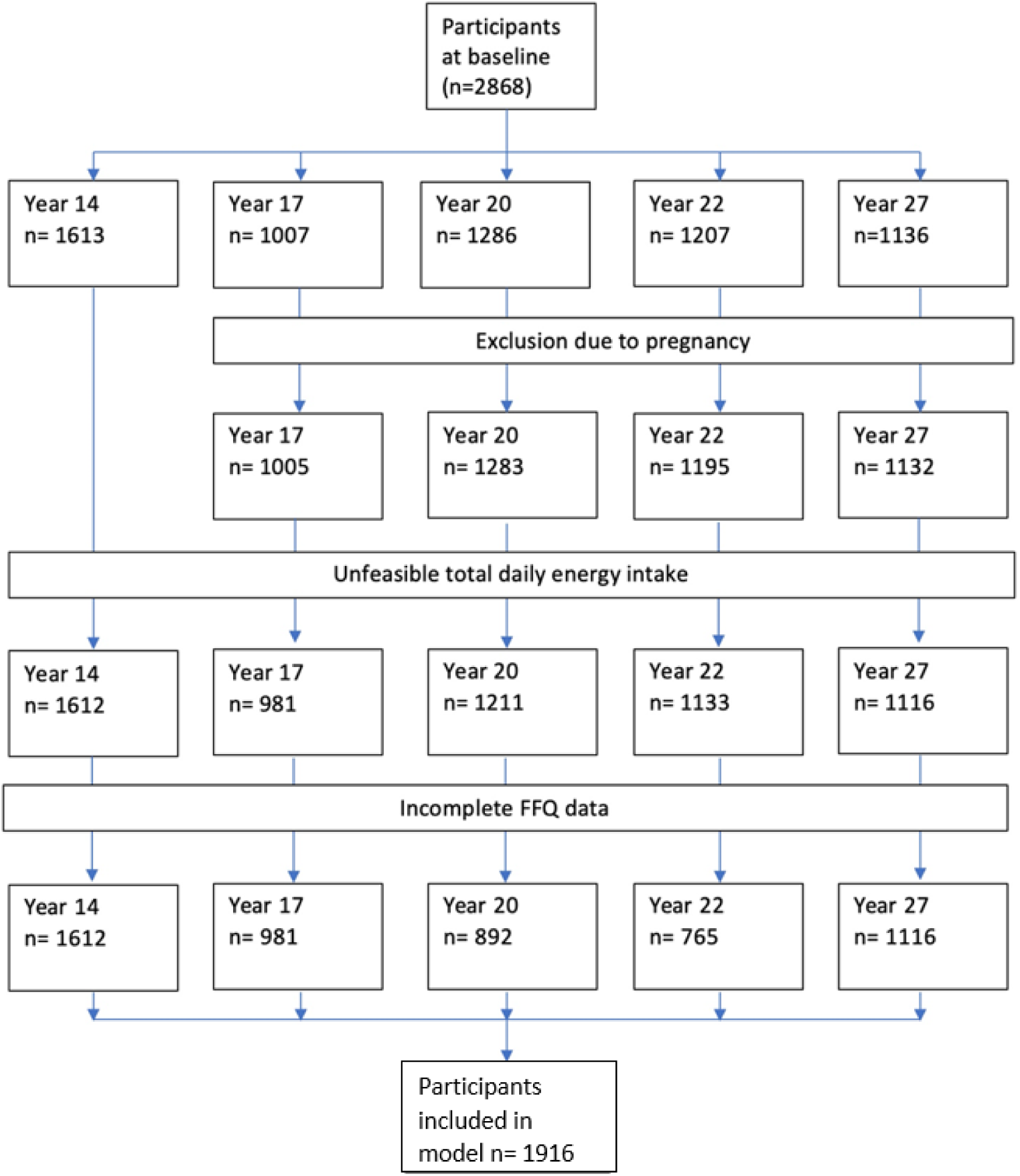
Number of participants included in study after exclusion criteria.

### 2.2 Diet measurement and diet quality

The primary outcome of this analysis was diet quality. Data on frequency of consumption and serving size of food items over the past year was self-reported using two distinct food frequency questionnaires (FFQs). The CSIRO FFQ (Baghurst and Record, 1984) at ages 14 and 17 and the Dietary Questionnaire for Epidemiological Studies (DQESV2) FFQ (Giles GG, Ireland PD., 1996) at ages 20, 22 and 27. FFQ data were converted to grams per day by the Cancer Council, Victoria, accounting for seasonal variation and portion size (Baghurst and Record, 1984; Giles GG, Ireland PD., 1996). In order to calculate diet quality scores, food intakes were first standardised to 2000kcal per day using the residual method (Willett, Howe and Kushi, 1997). They were then converted to servings per day at the level of food types (e.g. cheese, milk) using the Australian Nutritional Guidelines (Eat For Health, 2013) and subsequently aggregated into food groups (e.g. dairy).

Participants were assigned a Dietary Approaches to Stop Hypertension (DASH) diet quality score according to the system developed by Gunther and coauthors (Günther et al., 2009) based on the intake of eight different food groups. Three food groups (fats, sweets, meat, fish and eggs) were ‘moderation’ food groups, for which participants receive a lower score the more they consume above a set threshold. Five food groups (fruit, vegetables, dairy, nuts seeds and legumes) were ‘adequacy’ food groups for which a higher intake receives a higher score. Dairy intake was split into total dairy and proportion low-fat dairy coded as moderation and adequacy respectively. Similarly, Grain consumption was split into total grains, and proportion wholegrains (supplementary table 1). Intakes between the maximum and minimum thresholds were scored proportionally based on quantity consumed.

The Raine Study FFQs captured only a small number of foods that could be classified as ‘fats and oils’ as such underestimating intakes of this food group. To differentiate individuals based on fats consumption, the threshold for a maximum score for this food group was set as the sample median (across all time points) and the minimum score attributed to twice the median value.

### 2.3 Exposures

Parental SEP was assessed based on maternal education and household income. Level of maternal education was reported by the mother at birth as highest school year completed and highest qualification reached. This was categorised as: <y11: left school once legally allowed; y11-y12: left school after year 11 or completed school (year 12); or >y12: studied at a level beyond year twelve. Household income was measured in the years 2003 to 2005 when participants were age 14 and categorised into bands of annual income: $8000-30,000; $30,001-50,000; $50,001-70,000; $70,001-104,000; >$104,000. Individual SEP was assessed based on highest school year reached, self-reported at ages 17, 20, 22 and 27 years as school year 10 (typically age 15-16 years), year 11 (age 16-17 years), year 12 (age 17-18 years), and other (i.e. did not complete year 10). Individuals were categorised as: left school before completing school year 11; left school before completing school year 12 and left school after completing year 12 (the highest possible school year) based on their highest responses at ages 20, 22 and 27 years. For individuals who had left school by age 17, their response at this age was also used.

### 2.4 Covariates

Participant sex and maternal ethnicity were reported by the mother at birth of the participant. Participant age was self-reported at each recall of the cohort. Excessive alcohol consumption (had six or more drinks at one time/ so much that they vomited) at age 14 was self-reported as: never; once; or more than once. Overall health at age 14 was reported by parents as either: poor, so-so, ok, or excellent. Paternal education level was recorded at birth as per maternal education (above). Parental economic activity was recorded when participants were age 14 and categorised as: inactive (unemployed and not seeking employment); seeking work; paid work; unpaid work; and studying. Parental job skill level was categorised according to the Australia and New Zealand standard classification of occupations (ANZSCO 2022) with individuals assigned a number between 1 (highest level of skill and experience) and 5 (lowest level) at the sub-major level. Where the sub-major level comprised of two or three skill levels the higher and middle values were chosen respectively. Non-working parents were classed as economically inactive.

### 2.5 Missing data

Missing outcome data were handled in the growth mixture model using full information maximum likelihood (FIML). FIML produces unbiased estimates provided missing data are missing at random (MAR) or missing completely at random (MCAR) (Proust-Lima, Philipps and Liquet, 2015). Missing dietary data were considered to be MAR or MCAR, as the probability of missingness is unlikely to be influenced by diet quality.

Missing exposure and covariate data were handled using multiple imputation. In the exposure variables, data was missing in maximum school level by age 20 (21%), maternal education level (2%) and household income (13%). Missing exposure data stemmed primarily from individuals not responding to follow-up at the relevant age. Non-response was unlikely to be dependent on the value of the missing data and therefore exposure data were likely missing at random (MAR). Missing data were imputed using all variables and the outcome of the regression model (as per (Austin et al., 2021a)) and data on overall health, smoking status, alcohol consumption, and individual education level at later waves. Highest education level at age 22 and highest school year completed at age 22 were removed due to collinearity. 50 imputations were used based on the high level of missing data in some covariates (e.g. 37% missing in excessive alcohol consumption) (Austin et al., 2021b).

### 2.6 Analysis

Dietary trajectories were modelled using a growth mixture model (GMM). GMM is a type of latent trajectory analysis which offers additional flexibility over other methods (e.g. group-based trajectory models) by including random effects within diet quality trajectories to allow for between-individual variation within subgroups (Nagin, 2010). GMMs were fit using 100 random parameter starting points to reduce the likelihood of parameter estimates reaching a local maximum likelihood (Proust-Lima, Philipps and Liquet, 2015). All models included random intercept and random slope parameters to allow for between-person variation in both diet quality at age 14 and the change in diet quality over time. The distribution of random effects was allowed to vary by diet quality trajectory because trajectory groups were expected to differ in their homogeneity. Models were fit without covariates present to avoid making ex ante decisions regarding which factors shape dietary trajectories (as per (Nguena Nguefack et al., 2020). We did include the FFQ type used in dietary assessment as a covariate in our trajectory analysis to adjust for any differences in dietary reporting across the two FFQs. Participants were assigned to a diet quality trajectory according to their posterior probability for group membership.

Models were tested for each of linear, quadratic and cubic trajectory shapes. Additional diet quality trajectories were added and model fit compared across a suite of model fit indices, including Bayesian Information Criteria (BIC) and Akaike Information Criteria (AIC). Where model fit criteria were not in agreement, the Adjusted Lo-Mendell-Rubin likelihood ratio test (Lo, Mendell and Rubin, 2001) was used to decide between two models. To ensure that diet quality trajectories reflected real world subgroups, rather than just those who did not fit in other diet quality trajectories, models which included a diet quality trajectory containing less than 5% of the population were discarded (Nagin, 2005).

Mean food group intakes (servings/day) at each wave were described and compared pairwise between each diet quality trajectory and the reference trajectory using a two-tailed t-test. The characteristics of participants in each diet-quality trajectory (see exposure variables) were summarised using the raw (non-imputed) dataset as n(%) for each category. All exposure variables were categorical and thus compared pairwise between the reference trajectory and the other two diet quality trajectories using a chi-square test. Multinomial logistic regression was used to quantify the odds of membership in a dietary trajectory based on maternal education; household income, subsequently adjusted for other parental SEP indicators; and highest completed school year, adjusted for individual characteristics and health (overall health, and excessive alcohol consumption) at age 14 and then subsequently parental SEP (maternal and paternal education level, job skill level of primary carer and their partner and household income). All models were adjusted for participant age at each wave, participant sex and maternal ethnicity. We tested for moderation of associations by sex but found no consistent interaction effects across the exposures tested. See supplementary figures 1-3 for directed acyclic graphs for each model.

All analysis was performed using R version 4.3.3. The lcmm package was used to fit the GMM (Proust-Lima, Philipps and Liquet, 2015) and the nnet package was used for the multinomial regression model (Ripley, Venables and Ripley, 2016).

### 2.7 Ethics approval and consent to participate

Ethical approval was received from the Cambridge Psychology Research Ethics Committee, Application No: PRE.2022.062, updated 1st February 2024. The Raine Study adheres to the Australian National Health and Medical Research Council’s Guidelines for Ethical Conduct in Human Research (2018) and has received ethical approval from the King Edward Memorial Hospital for Women, the Princess Margaret Hospital for Children and The University of Western Australia, in Perth, Western Australia. Primary caregivers (at ages 14 and 17) and participants (at ages 20, 22, and 27) provided informed written consent.

## 3 Results

### 3.1 Diet quality trajectories

A quadratic model with three diet quality trajectories was selected as the best fitting model for the change in DASH score over time (see supplementary table 2). The two-class quadratic model had similar model fit criteria according to AIC and BIC, however an adjusted Lo-Mendel-Rubin likelihood ratio test indicated that model fit improved with the additional class (p-value <0.001). The three classes (figure 2), characterised by their shape and intercept, are as follows: High diet quality trajectory (n = 312, 16%): high mean DASH score throughout follow up with a shallow positive increase with age. Low-to-High (n = 811, 42%): lowest mean DASH score at mean age 14 but displays a steep increase in DASH score over time before plateauing between age 22 and 27. Consistently low (n = 793, 41%): low mean DASH score at mean age 14 decreasing up to age 20 before increasing. The low-to-high class was designated the reference class as it was the largest class.

**Fig. 2.**
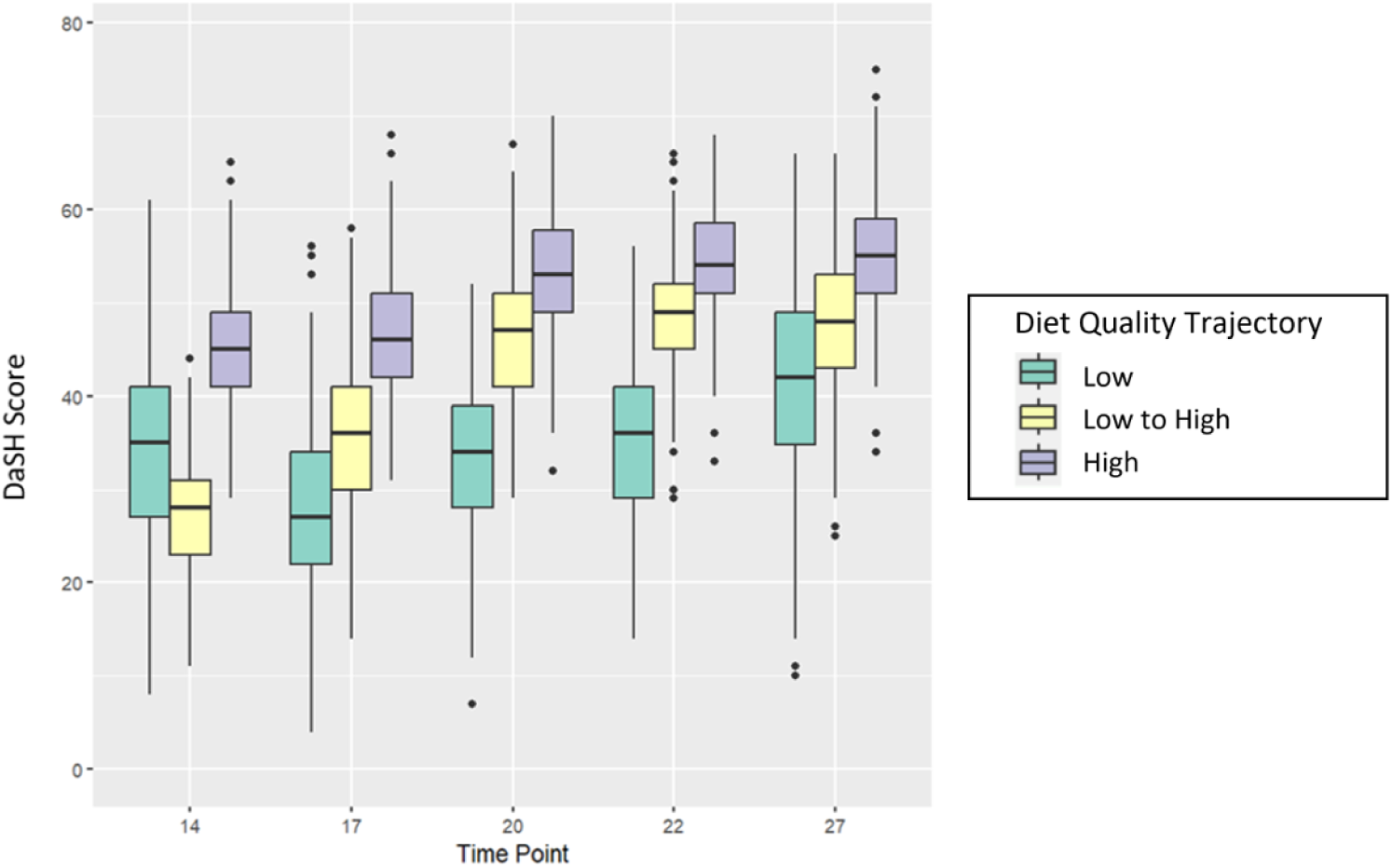
DASH diet quality score at each age of data collection across the three diet quality trajectories: Low quality (n= 793 (41%)), low to high (n= 811 (42%), and high quality (n= 312 (16%)). and Box and whisker plots show median, quartiles and range of the DASH score at each age.

### 3.2 Food group intakes

For all diet quality trajectories, mean fruit consumption was consistently lower than the maximum score threshold (*≥*4 servings/day) and declined with age (figure 3). Vegetable and nuts and legumes intakes were mostly below the maximum score threshold but improved with age, particularly between age 22 and 27. Total grain consumption decreased with age whilst the share of wholegrain increased. Dairy consumption decreased with age, with most choosing low-fat options at all ages. By age 27, there were many low or non-consumers of dairy who received a minimum score for both total and low-fat dairy. Meat fish and egg consumption followed a U-shaped pattern with median consumption at its lowest level at age 20. Fat consumption decreased over time. Median sweets (SSBs and confectionary) consumption started high but decreased sharply after age 17 (figure 3, Supplementary data table 3: food group intake by diet quality trajectory).

**Fig. 3.**
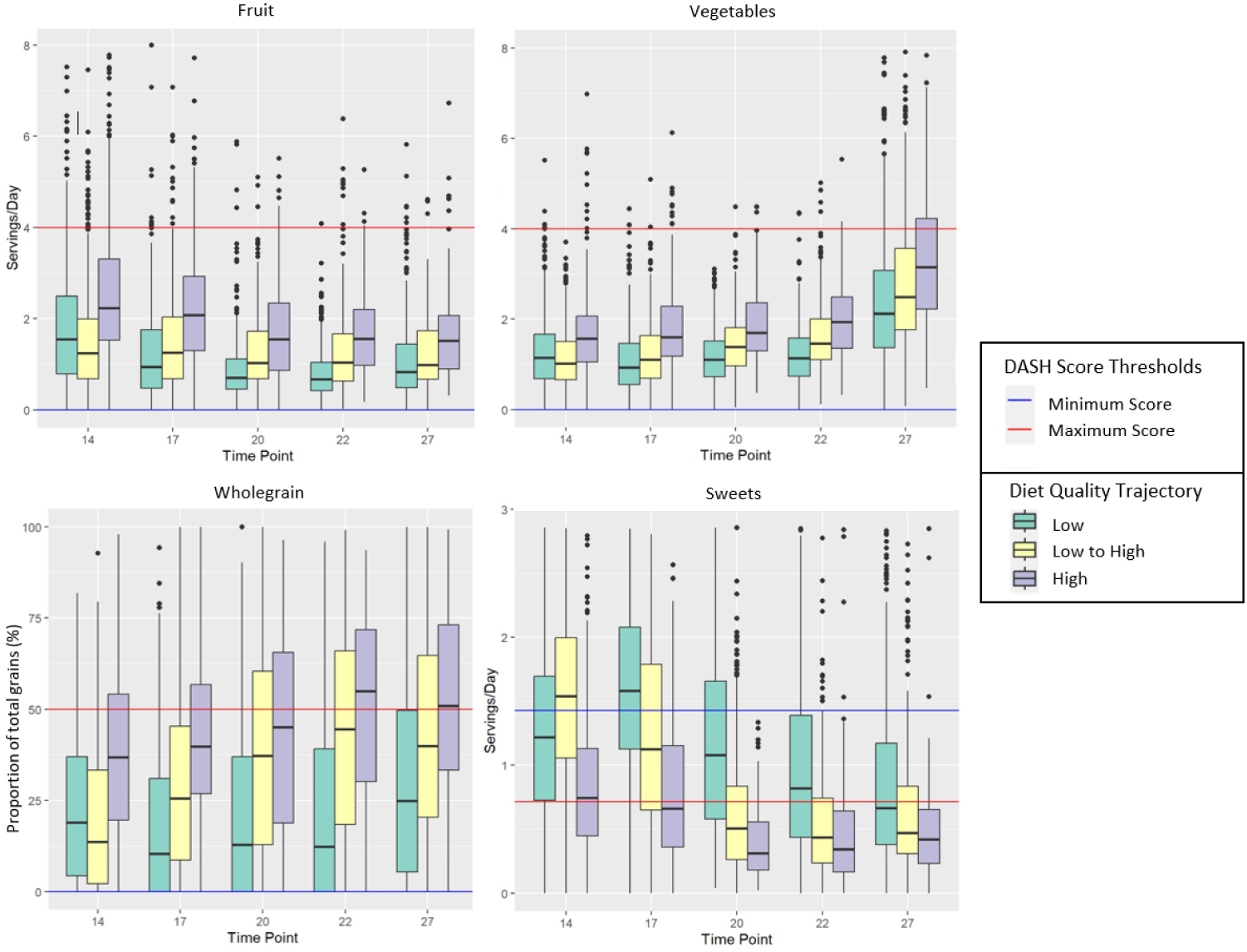
Intakes of fruit, vegetables, wholegrains and sweets at each wave of data collection across the 3 diet quality trajectories. Box and whisker plots show median, quartiles and range of the DASH score at each age.

Compared to the low to high diet quality trajectory, the high diet quality trajectory scored consistently higher for intakes of nuts and legumes, vegetables, fruit, and wholegrain cereals. The high diet quality trajectory scored on average higher for sweets consumption at ages 14 and 17, but similarly from age 20 onwards. The two classes scored similarly in all other food groups (figure 3). Compared to the low to high diet quality trajectory, the low diet quality trajectory scored similarly in all food groups at age 14. From age 17 onwards the low diet quality trajectory scored consistently lower for intakes of vegetables, nuts and legumes, meat fish and eggs, wholegrain cereals and sweets. The low diet quality trajectory scored lower on fruit intake up to age 27 where they scored similarly (figure 3).

### 3.3 Socioedemographic variables and diet quality trajectories

The low diet quality trajectory had the greatest proportion of males (59.3%) and the high diet quality trajectory the lowest (35.3%) (Table 1). Ethnicity differed little between diet quality trajectory, with all classes predominantly “Caucasian” (>90%) followed by Chinese (4%) and Indian (1%-3%). There were few participants of Aboriginal or Polynesian descent (*≤*1%).

**Table 1.**
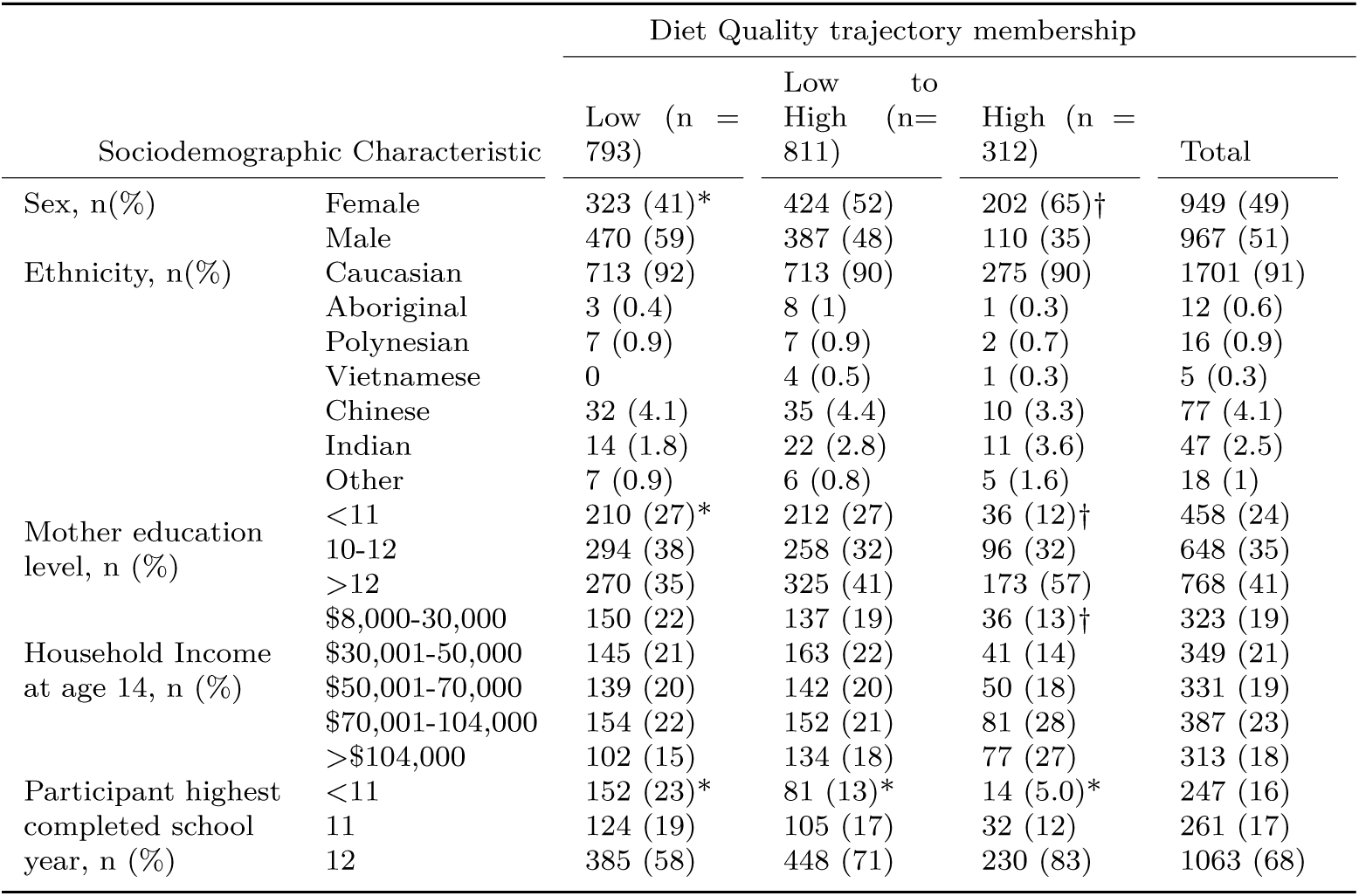
Sociodemographic characteristics of the population following each diet quality trajectory. * indicates a p-value of *<*0.05 for chi-square test between low and low to high diet quality trajectories. † indicates a p-value *<*0.05 for chi-square test between low to high and high diet quality trajectories.

SEP characteristics also differed by latent class; Compared to the low- to-high diet quality trajectory, members of the low diet quality trajectory had lower maternal education and a lower proportion completed school (y12). The opposite was true for the high diet quality trajectory compared to the low- to-high diet quality trajectory. Further, a lower proportion of participants in the high diet quality trajectory lived in households in the lowest income band and a greater proportion in the highest income band than members of the low to high diet quality trajectory (table 1).

Having a mother who completed education beyond year 12 rather than left school between years 10-12 was negatively associated with membership in the low diet quality trajectory (OR = 0.72 [95% CI: 0.57, 0.91] (Table 2). In contrast, having a mother who left school before completing year 10 was negatively associated (OR = 0.47 [95% CI: 0.31, 0.71], and having a mother who completed education beyond year 12 positively associated (OR = 1.44 [95% CI: 1.07, 1.94] with membership in the high diet quality trajectory. After controlling for participant sex and age, being in the highest band of household income (>$104,000/year) compared to the lowest band of income (<$30,000/year) was negatively associated with membership in the low diet quality trajectory (OR=0.65 [95% CI: 0.50, 0.85]. Being in one of the highest two bands of income ($70,001->$104,000 and >$104,000) was positively associated with membership in the high diet quality trajectory (OR = 1.92 [95% CI: 1.46, 2.51] and 2.08 [95% CI: 1.57, 2.76] respectively). These associations between household income and class membership attenuated only slightly after controlling for other parental SEP indicators.

**Table 2.** Associations of parental and individual socioeconomic position with membership of different diet quality trajectories. Odds ratios are presented for following the low or high diet quality trajectory, compared to the low-to-high diet quality trajectory (reference class). Associations with parental SEP indicators and participant highest completed school year were assessed in separate models. See supplementary figures 1-3 for directed acyclic graphs for each model.

| Variable |  | Odds Ratio for Diet Quality Trajectory Membership, OR (95% CI) |  |  |  |
| --- | --- | --- | --- | --- | --- |
|  |  | Low diet quality |  | High diet quality |  |
| Parental Characteristics |  |  |  |  |  |
|  |  | Adjusted for participant sex, age, and ethnicity | Adjusted for parental education and job skill level<sup>1</sup>, and participant sex and age | Adjusted for participant sex, age, and ethnicity | Adjusted for parental education and job skill level<sup>1</sup>, and participant sex and age |
| Maternal Education level | 10 | 0.88 (0.68-1.14) |  | 0.47 (0.31-0.71) |  |
|  | 10-12 | 1 |  | 1 |  |
|  | >12 | 0.72 (0.57-0.91) |  | 1.44 (1.07-1.94) |  |
| Household income (age 14y) | <\$30,000 | 1 | 1 | 1 | 1 |
| | \$30,000-50,000 | 0.83 (0.62-1.10) | 0.81 (0.62-1.04) | 0.92 (0.67-1.26) | 0.98 (0.68-1.40) |
| | \$50,001-70,000 | 0.87 (0.66-1.16) | 0.83 (0.64-1.07) | 1.27 (0.94-1.72) | 1.39 (1.01-1.90) |
| | \$70,001-104,000 | 0.89 (0.67-1.17) | 0.87 (0.68-1.12) | 1.92 (1.46-2.51) | 1.76 (1.34-2.32) |
| | >\$104,000 | 0.65 (0.48-0.88) | 0.65 (0.50-0.85) | 2.08 (1.57-2.76) | 1.56 (1.17-2.08) |
| Individual Characteristics |  |  |  |  |  |
|  |  | Adjusted for participant sex, age, ethnicity, and health<sup>2</sup> | Additionally Adjusted for parental SEP1 | Adjusted for participant sex, age, ethnicity, and health<sup>2</sup> | Additionally Adjusted for parental SEP1 |
| Highest completed school year | <11 | 1.76 (1.34-2.39) | 1.59 (1.13-2.23) | 0.43 (0.25-0.75) | 0.56 (0.31-1.03) |
|  | 11 | 1.23 (0.92-1.65) | 1.1 (0.80-1.52) | 0.69 (0.45-1.06) | 0.80 (0.50-1.28) |
|  | 12 | 1 | 1 | 1 | 1 |

<sup>1</sup>Parental education includes both maternal and paternal education level, and job skill level both primary carer and partner of primary carer job skill level.
<sup>2</sup>Individual health age 14 included as parent-reported overall health, and excessive alcohol consumption.

Compared to completing school (finishing year 12), leaving school before completing year 11 was significantly positively associated with membership in the low diet quality trajectory (OR = 1.76 [95% CI: 1.34, 2.39]) independent of individual sociodemographic characteristics. This association was attenuated only slightly upon additionally adjusting for parental SEP (OR = 1.59 [95% CI: 1.13, 2.23] In contrast, leaving school before completing year 11 was negatively associated with membership in the high diet quality trajectory (OR = 0.43 [95% CI: 0.25, 0.75]. However, this association became non-significant upon adjustment for parental SEP.

## 4 Discussion

### 4.1 Summary of main findings

This study identified three distinct diet quality trajectories within the Raine Study Gen2 population, a high diet quality trajectory (16% of participants), a low to high diet quality trajectory (42%) and a low diet quality trajectory (41%). Those following the high diet quality trajectory consumed a consistently higher quality diet from the age of 14 to 27yrs. Membership of this trajectory was associated with higher maternal education and higher parental household income. Participants in the two remaining diet quality trajectories had similar lower diet quality scores at age 14. However, by age 17 these two trajectories had diverged. Diet quality in the low to high diet quality trajectory improved with age whilst that of the low diet quality trajectory first decreased before improving more gradually. The low diet quality trajectory had a high proportion of male participants (59%) whilst a greater proportion of females than males followed the high diet quality trajectory (65%). Membership of the low diet quality trajectory was associated with lower maternal education a lower parental household income. Membership of this trajectory was also associated with participants leaving school early, before completing school year 11, independent of parental socioeconomic position.

### 4.2 Strengths and limitations

A key strength of this study was the use of latent trajectory analysis to study diet trajectories across 5 waves, which allowed the study to capture the change in diet quality across the period of adolescence and early adulthood. This improved our capacity to determine the shape of trajectories and provided information into diet quality and its determinants across an understudied period of the life course. We assessed diet quality using the DASH score which has strong evidence for association with health outcomes (Appel Lawrence J. et al., 1997), including in adolescents (Moore et al., 2009; Asghari et al., 2016). The use of the DASH score also allowed us to identify which food groups drove the difference in diet quality between trajectories. In our analyses we were able to control for a number of potential confounders and model the effects of both parental and individual socioeconomic status. This improved the study’s capacity for causal inference and allowed us to determine the effect of individual socioeconomic position independent of parental socioeconomic position on diet trajectories. Gen2 of the Raine Study have been shown to be representative of the Western Australia population, with the exception of a higher loss to follow up of participants Aboriginal and Torres Strait Islander ethnicity over time compared to other ethnicities (Straker et al., 2017; White et al., 2017).

This study did however have a number of limitations. In line with many studies, use of an age-appropriate FFQ in adolescence meant that the FFQ used to collect dietary data changed between ages 17 and age 20. We accounted for this in our analysis by including a dummy-variable for FFQ type, and we believe this will have had limited impact on the trajectory classification. The Raine Study is an observational study; as such, it was not possible to control for secular changes in diet quality during the study period or for all possible confounders. Finally, the latent trajectory model chosen in this study had a relatively low entropy value. The indicates that the model had difficulty classifying individuals into distinct diet quality trajectories. This may stem from the DASH diet score used to assign participants a diet quality score or the nature of the dietary data collected by the Raine Study.

### 4.3 Comparison with previous research and implications of the findings

Other longitudinal studies assessing diet quality during adolescence and early adulthood have identified distinct dietary trajectories, however the exact number and shape of these trajectories vary (Appannah et al., 2021; Doggui et al., 2021; Dalrymple et al., 2022). These differences may stem from methodological differences and temporal and socioenvironmental differences between study populations. Our findings are in line with studies in the UK and the USA that have shown a general improvement in diet quality from late adolescence into adulthood (Winpenny et al., 2018; Tao et al., 2024).

Diet quality in each of the three classes changed substantially over time. Cross-sectional methods would not capture these temporal changes and may miscategorise an individual’s diet quality depending on the age chosen. For example, despite having a similar quality of diet at age 14, members of the low diet quality trajectory were exposed to a substantially worse quality diet between ages 14 and 27 than those in the low-to-high class. This inequality in diet quality remained at the end of follow-up, suggesting it may continue into adulthood. The observed inequalities in diet quality may in turn lead to inequalities in chronic disease risk between the diet trajectory classes. This could be explored further in future research.

Our findings were in accordance with longitudinal studies which have shown that dietary trajectory membership during adolescence and early adulthood is associated with parental socioeconomic position (Dalrymple et al., 2022). Our findings are also consistent with previous studies which have shown that diet quality and diet quality trajectories differ by sex during adolescence and early adulthood, with males having a worse quality diet than females (Doggui et al., 2021, Winpenny et al., 2018, Tao et al 2024). Indeed, Appannah and coauthors (2021) found that in the Raine Study cohort males living in a lower income household were more likely to follow a trajectory with an increasing Western Diet score over time, which is defined by an unhealthy dietary pattern, than males and those in higher income households. Further, Appannah and coauthors found that a lower maternal education level was associated with a lower Healthy Diet score during adolescence and early adulthood.

Our study added to this existing knowledge, showing that different diet trajectories differed in their association with individual, in addition to parental socioeconomic position. This has important implications for the development of the socioeconomic inequalities in diet and cardiovascular health which have been observed later in life (Darmon and Drewnowski, 2008, de Mistral and Stringhini 2017). While previous research has shown early adulthood to be a key period for development of dietary and cardiovascular inequalities (Winpenny et al., 2021) there has previously been little insight into when and how these inequalities develop.

A key finding of our study was that individuals who left school before year 11 were at particular risk of a low diet quality trajectory during early adulthood. Specific food groups which drove this included high consumption of confectionary and sugar sweetened beverages, and a low consumption of fruits and vegetables (see supplementary table 3). There is little research into the effect of leaving school on diet quality, however one longitudinal study has shown that leaving education at any age was associated with increased consumption of SSBs and confectionary in a Norwegian population (Winpenny et al., 2018). A second longitudinal study found that leaving education, particularly at a younger age, was associated with decreases in diet quality (Tao et al., 2024).

Similar to the ‘structured days’ hypothesis, leaving school could influence diet quality due to the loss of the structure that students have at school (Brazendale et al., 2017). Further, school leavers who enter the workplace may be exposed to factors which negatively impact diet quality. For example, increased time constraints due to work commitments (Holley et al., 2016; Munt, Partridge and Allman-Farinelli, 2017) and increased proximity to unhealthy foods including take-aways and sugar sweetened beverages (Watts et al., 2016). The impact of leaving school early may be an important factor in the development of the widely observed dietary inequalities in adulthood according to maximum education level. Future research should explore the reasons for poor diet quality in those who leave education at a young age and interventions to improve dietary quality in this group should be considered.

## 5 Conclusion

This study identified three distinct diet quality trajectories present during adolescence and early adulthood. The three trajectories not only differed in their diet quality but also in their associations with both individual and parental socioeconomic position. In particular, individuals who leave school early show persistently low diet quality whilst others improve their diet quality as they move into adulthood. Interventions need to consider the distinct diet quality trajectories followed by different subgroups of the population. Our findings indicate that interventions to improve diet quality in early adulthood are needed amongst those who leave education at a young age.

## Data Availability

Information regarding accessing data from the
Raine Study can be found at rainestudy.org.au

## Acknowledgements

We would like to acknowledge the Raine Study participants and their families for their ongoing participation in the study and the Raine Study team for study co-ordination and data collection. We also thank the National Health and Medical Research Council of Australia (NHMRC) and the Raine Medical Research Foundation for their long term contribution to funding the study over the last 30 years.

## 6 Declarations

- Consent for publication Not required
- Availability of data and materials Information regarding accessing data from the Raine Study can be found at rainestudy.org.au
- Funding OR is jointly funded by the Wellcome Trust Doctoral Training Centre in Public Health Economics and Decision Science [108903] and the University of Sheffield. EW is funded by the UK Medical Research Council MR/T010576/1. TB is funded by the Cambridge MRC Doctoral Training Partnership (grant number MR/N013433/1) and the Elizabeth McDowell Studentship at Newnham College, Cambridge. The core management of the Raine Study is funded by The University of Western Australia, Curtin University, Telethon Kids Institute, Women and Infants Research Foundation, Edith Cowan University, Murdoch University and The University of Notre Dame Australia.
- CRediT Author contribution OR: conceptualisation, formal analysis, visualisation, writing (orginal draft), writing (review and editing); TB, TM, LB: writing (review and editing); EW: conceptualisation, writing (review and editing), supervision.
- Competing interests No competing interests.

---

**Table 1.**
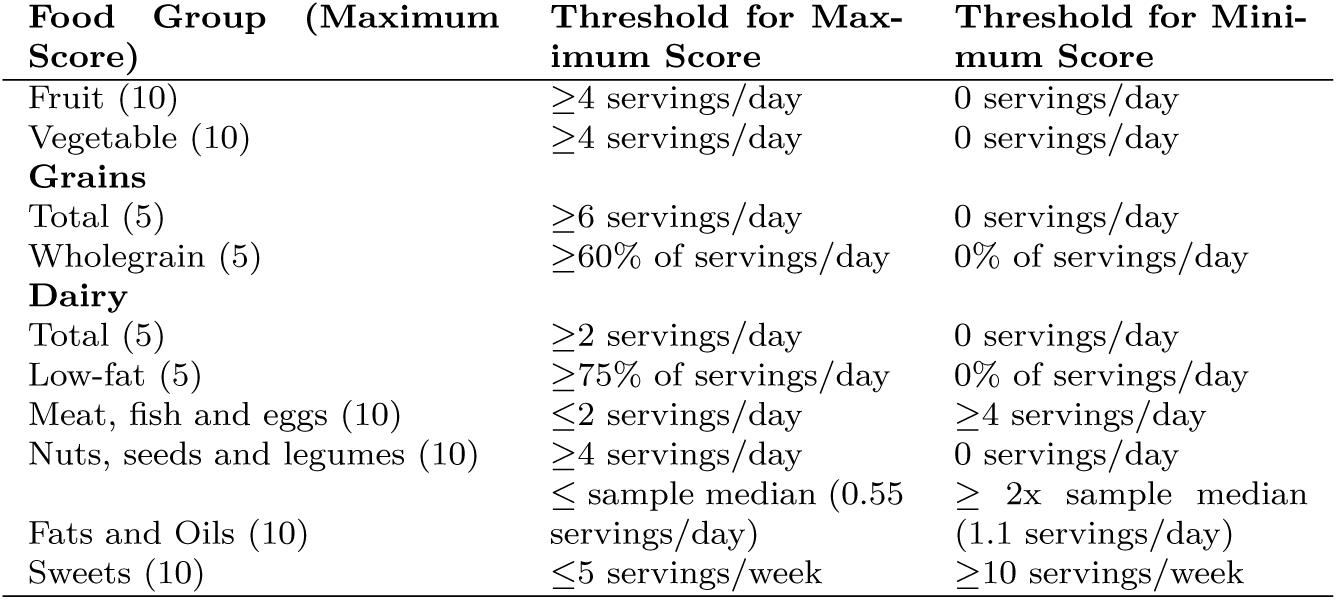
DASH scoring algorithm used to calculate diet quality.

**Fig. 1.**
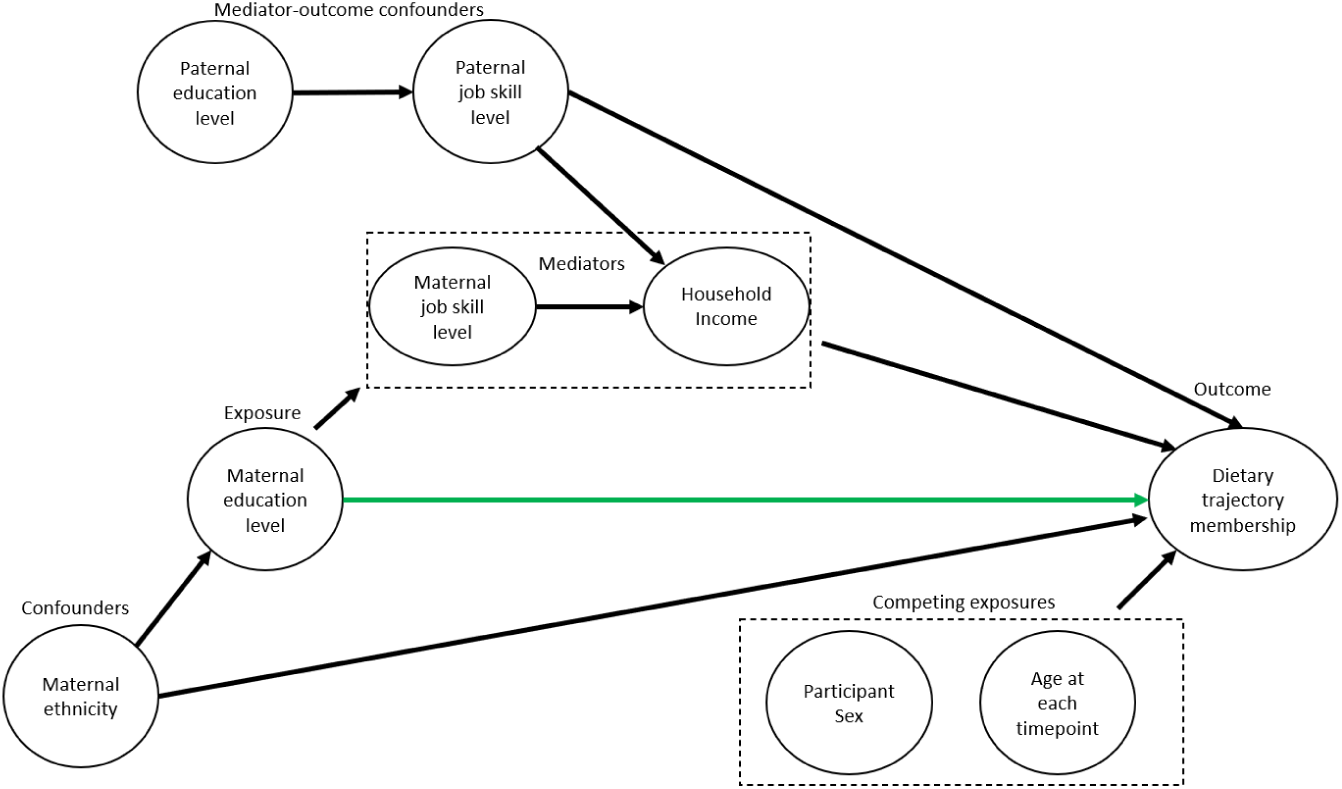
Directed Acyclic Graph for the model investigating the association between maternal education level and diet quality trajectory. Green line shows association of interest.

**Fig. 2.**
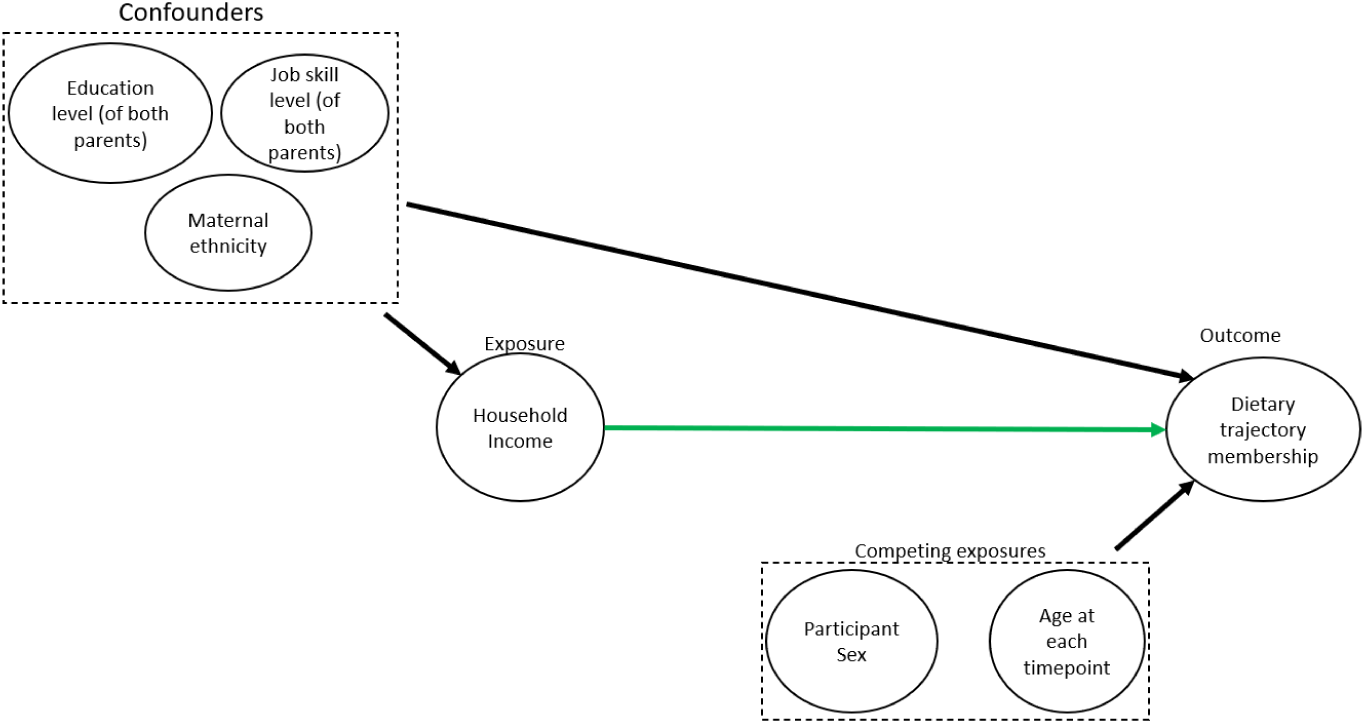
Directed Acyclic Graph for the model investigating the association between household income and diet quality trajectory. Green line shows association of interest.

**Table 2.** Model fit criteria for growth mixture models. * P-values for a lo-mendel-rubin adjusted likelihood ratio test between model with n classes and alternative model with the same parameters but n-1 classes, testing the null hypothesis that inclusion additional class does not improve model fit.

| Number of classes | Maximum Likelihood | AIC | BIC | Entropy | Lo-Mendel-Rubin LRT (p-value)* | Class size (%) |  |  |  |
| --- | --- | --- | --- | --- | --- | --- | --- | --- | --- |
|  |  |  |  |  |  | Class 1 | Class 2 | Class 3 | Class 4 |
| Linear shape |  |  |  |  |  |  |  |  |  |
| 1 | -19541 | 39096 | 39135 | 1 | - | 100 | - | - | - |
| 2 | -19514 | 39050 | 39111 | 0.66 | <0.001 | 5 | 95 | - | - |
| 3 | -19506 | 39042 | 39126 | 0.61 | 0.005 | 4 | 90 | 6 | - |
| 4 | -19506 | 39044 | 39149 | 0.40 | 0.224 | 3 | 40 | 42 | 14 |
| Quadratic shape |  |  |  |  |  |  |  |  |  |
| 1 | -19540 | 39096 | 39140 | 1 | - | 100 | - | - | - |
| 2 | -19468 | 38962 | 39034 | 0.28 | <0.001 | 31 | 69 | - | - |
| 3 | -19456 | 38947 | 39047 | 0.36 | <0.001 | 41 | 42 | 16 | - |
| 4 |  |  |  |  | Did not converge |  |  |  |  |
| Cubic Shape |  |  |  |  |  |  |  |  |  |
| 1 | -19538 | 39095 | 39145 | 1 | - | 100 | - | - | - |
| 2 | -19466 | 38962 | 39046 | 0.28 | <0.001 | 70 | 30 | - | - |
| 3 | -19444 | 38930 | 39047 | 0.30 | <0.001 | 31 | 33 | 36 | - |
| 4 | -19430 | 38915 | 39065 | 0.35 | <0.001 | 20 | 25 | 43 | 12 |

**Table 3.**
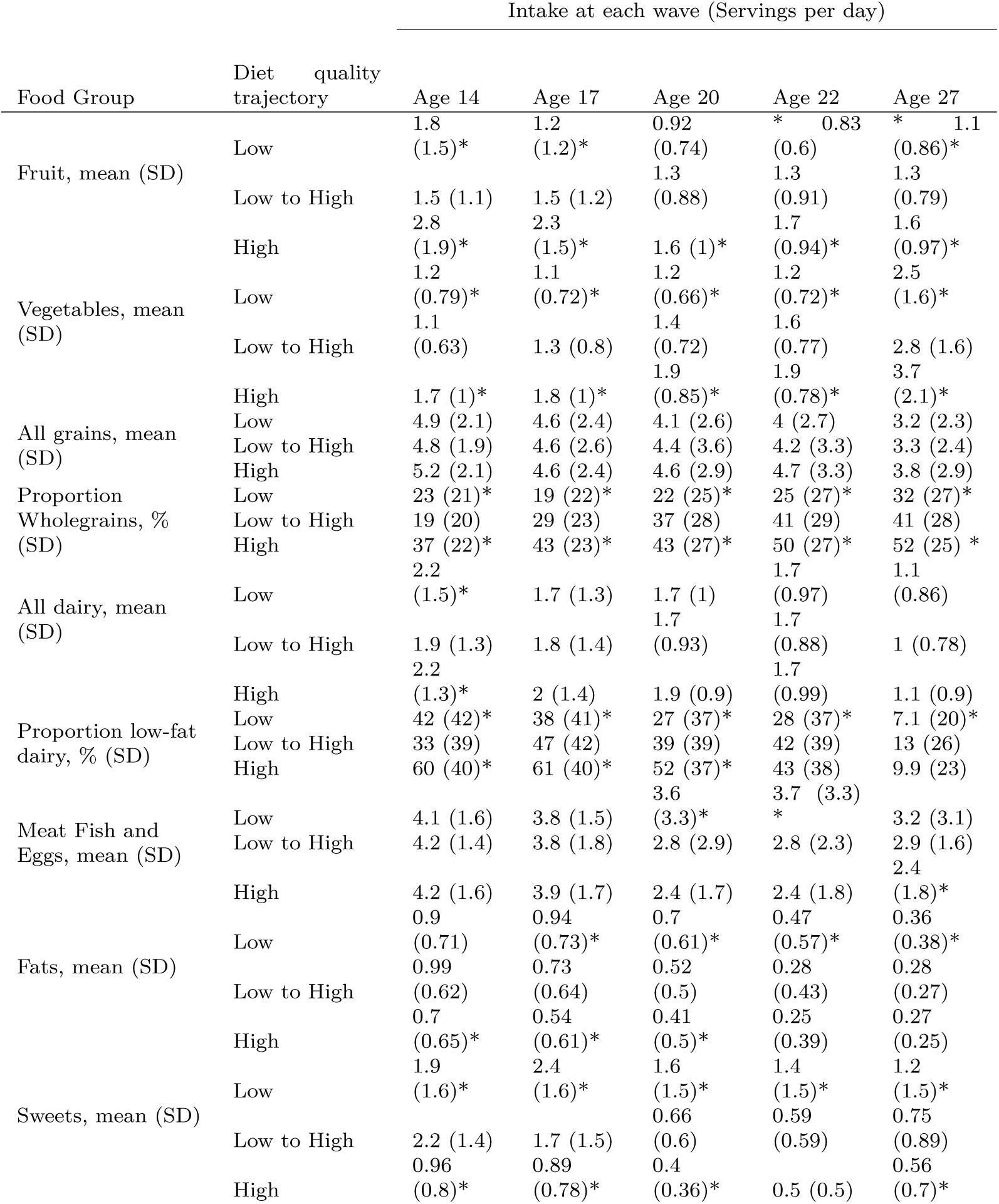
Intake of food groups by diet quality trajectory and wave. T-test conducted between intake at each wave between the low- and high-diet quality trajectories and the low-to-high class. * represents a corresponding p-value of ¡0.01.

**Fig. 3.**
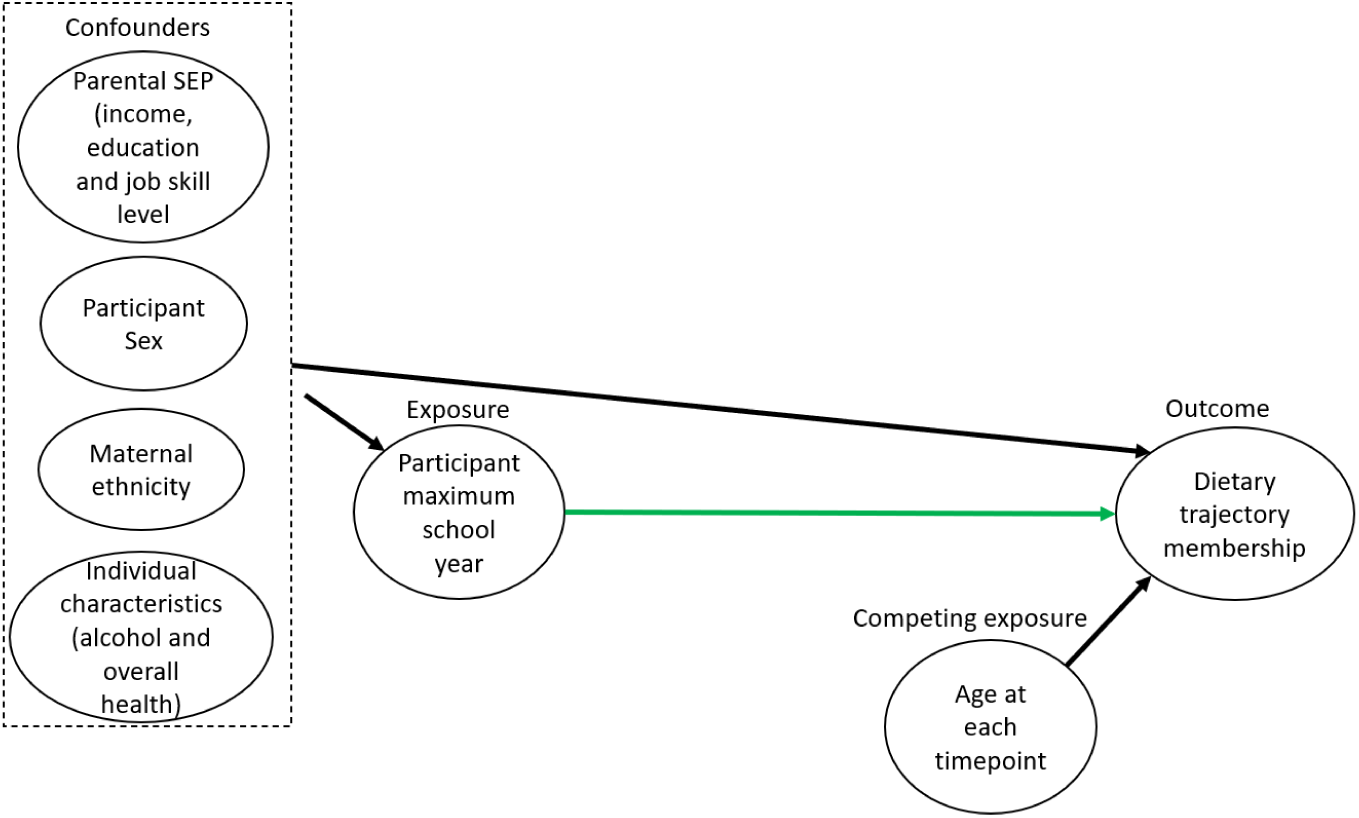
Directed Acyclic Graph for the model investigating the association between participant highest completed school year and diet quality trajectory. Green line shows association of interest.

## Notes

### Competing Interest Statement

The authors have declared no competing interest.

### Author Declarations

Psychology Research Ethics Committee of The University of Cambridge gave ethical approval for this work. Application No: PRE.2022.062 updated 1st February 2024

### Summary of Updates

Author list order changed to correct previous error.

